# A Comprehensive Study of Circulating Blood Linear RNA nominates CD55 and DLD as novel causal genes and early-stage biomarkers for Parkinson’s Disease

**DOI:** 10.1101/2025.06.20.25329948

**Authors:** Aleksandra Beric, Sarp Sahin, Santiago Sanchez, Zining Yang, Ravindra Kumar, Isabel Alfradique-Dunham, Jessie Sanford, Daniel Western, Bridget Phillips, John P. Budde, Richard J. Perrin, Paul T. Kotzbauer, Joel S. Perlmutter, Scott A. Norris, Carlos Cruchaga, Laura Ibanez

**Affiliations:** Department of Psychiatry, Washington University in Saint Louis School of Medicine; NeuroGenomics and Informatics Center, Washington University in Saint Louis School of Medicine; Department of Neurology, Washington University in Saint Louis School of Medicine; Department of Pathology & Immunology, Washington University in Saint Louis School of Medicine; Department of Radiology, Washington University in Saint Louis School of Medicine; Department of Neuroscience, Washington University in Saint Louis School of Medicine; Program of Physical Therapy, Washington University in Saint Louis School of Medicine; Department of Occupational Therapy, Washington University in Saint Louis School of Medicine; Hope Center for Neurological Disorders, Washington University in Saint Louis School of Medicine; The Charles F. and Joanne Knight Alzheimer Disease Research Center, Washington University in Saint Louis; Department of Genetics, Washington University in Saint Louis School of Medicine

## Abstract

We leveraged transcriptomic data from 4,343 participants from four independent datasets to robustly identify and annotate circulating PD-associated transcripts. We identified 296 differentially expressed transcripts, 28 of which were transcribed from known PD-associated loci. Further, we found a significant overlap between our findings and transcripts dysregulated in brain, as well as proteins differentially accumulated in CSF. Expression of the identified transcripts was affected by genetic background including ancestry and PD-related mutations, and nearly half of the identified transcripts were dysregulated before symptom onset. The differentially expressed transcripts were utilized to develop three predictive models that distinguished between PD and healthy controls with a ROC AUC of 0.727-0.733. The predictive models were capable of detecting PD transcriptomic signatures even before symptom onset. One transcript, DLD, showed particular promise as an early stage, minimally invasive PD biomarker that was differentially expressed in whole blood, brain and CSF. This transcript significantly related to PD in the eQTL analyses and in two of the three predictive models.

## INTRODUCTION

Parkinson’s disease (PD) is a progressive neurodegenerative disease characterized by motor (e.g. resting tremor, postural instability, bradykinesia), and non-motor features (e.g. cognitive decline, depression, pain).^1,2^ Affecting 2-3% of the population over 65, PD is the second-most common neurodegenerative disorder and its prevalence may double between 2015 and 2040.^3,4^ PD is pathologically defined by the accumulation of misfolded and abnormally phosphorylated alpha-synuclein (α-Syn) protein in Lewy bodies and neurites within the brain.^5^ This protein aggregation occurs alongside degeneration of dopamine-producing neurons in the substantia nigra, a critical brain region for movement control.^5^ Current therapeutic strategies focus on managing PD symptoms, but no current therapy prevents neuronal degeneration or slows disease progression. A deeper understanding of the underlying mechanisms driving PD pathogenesis is imperative to develop more effective treatments which could include gene-targeted therapies.

Blood transcriptomics offers a compelling approach to investigate PD pathogenesis. Compared to invasive tissue biopsies or expensive imaging procedures, blood draws are minimally invasive and readily available in most healthcare settings. Moreover, whole blood, which shares over 80% overlap with neuronal gene expression, holds potential for capturing meaningful molecular changes at the transcript level.^6–9^ Linear RNA, in particular, reflects actively transcribed genes within cells, providing a real-time snapshot of cellular processes. This dynamic view of cellular activity has been leveraged in previous PD transcriptomic studies to consistently identify disease-associated genes, transcripts, and enriched pathways.^10^ For instance, a study measuring differences in blood RNA expression data identified and validated eight genes associated with PD risk from a recent meta-analysis of genome-wide association studies (GWAS).^11,12^ Further supporting the utility of blood-based analysis, Irmady et al. identified 1,584 transcripts that were significantly dysregulated in PD blood and showed similar changes in expression within the caudate and putamen regions of the PD brain.^13^ Makarious et al. integrated whole blood RNA-seq data with clinical, demographic and genome sequencing data to develop a multi-modal model capable of predicting PD with 85.03% accuracy and 93.12% sensitivity, demonstrating that the blood transcriptome captures biologically significant changes that, combined with other data modalities can improve the accuracy of PD diagnosis.^14^ However, the generalizability of most of the transcriptomic findings has been limited by relatively small sample sizes and use of different methodologies.

The search for diagnostic biomarkers for PD has intensified in recent years, driven by the promise of earlier detection and intervention. While recent studies proposed some promising PD biomarker candidates, such as α-synuclein seed amplification assay^15^ and α-/β-synuclein ratio^16^, identification of non-invasive PD biomarkers remains paramount as it would greatly aid disease diagnosis and monitoring.

To address this gap in knowledge, we conducted a comprehensive analysis of whole blood transcriptomic profiles in the largest PD cohort to date. We have processed and meta-analyzed the transcriptomic profiles from four major studies: the Parkinson’s Disease Biomarkers Program (PDBP)^17^, the Parkinson’s Progression Markers Initiative (PPMI)^18^, the Fox Investigation for New Discovery of Biomarkers (BioFIND)^19^, and Washington University in St. Louis Movement Disorders Clinic cohort (WashU-MDC). Then, we performed pathway analyses and multi-omic data integration to understand the biological implication of our findings. We identified if the transcripts are dysregulated in brain and if the protein products are dysregulated in plasma and/or CSF of PD participants. Next, we examined whether those transcripts were encoded in known PD-associated genetic loci, identified cis-expression quantitative trait loci (eQTL) and performed Mendelian Randomization (MR) to infer causality. Finally, we assessed the predictive power of the blood transcriptome by leveraging it to build preliminary biomarkers for disease detection and follow-up.

## METHODS

### Study design

We leveraged four independent PD whole blood RNAseq datasets: PDBP^17^ (N=1,604), PPMI^18^ (N=1,943), BioFIND^19^ (N=213), and WashU-MDC^20^ (N=583; Figure1). We performed differential expression (DE) analyses in each cohort separately and then combined the findings from all four cohorts via meta-analysis. Only transcripts with the same direction of effect in all four cohorts were assessed in the meta-analysis, and those with multiple-test corrected p-values lower than 0.05 in the meta-analysis were considered DE. Next, we biologically contextualized our findings through pathway analyses and integration with an in-house brain transcriptomic and publicly available plasma and CSF proteomic datasets. We also leveraged the available PD GWAS resources to verify whether any of the DE transcripts originated from known PD-risk loci. For all significant transcripts, we identified the eQTLs and tested causality using MR. Further, we evaluated the effect that ancestry and PD-related mutations had on the whole blood transcriptomic landscape. Then, we tested whether there is detectable transcript dysregulation in participants without symptoms, but at risk of developing PD. We explored clinical importance of our findings by investigating the impact of medication on the expression of the identified transcripts, as well as the relationship of the DE transcripts with UPDRS-III and MoCA scores, measures of motor symptom and dementia severity, respectively. Finally, we leveraged our findings to develop predictive models capable of distinguishing between PD and healthy control participants. This study was approved by the Washington University in Saint Louis Institutional Review Board (IRB IDs: 201701124 and 202004010).

### Study population

PDBP and PPMI enrolled participants who were followed longitudinally with clinical assessments and biospecimen collection every six months. Blood RNA-seq data was collected every six months in PDBP, and every six months during the first year followed by annually thereafter in PPMI. Instead of using baseline as previous publications^11,21^ have done for cross-sectional analyses, we selected the last assessment of each participant in an attempt to maximize clinical and molecular differences between cases and controls, and harmonize with the BioFIND and WashU-MDC cohorts inclusion criteria. BioFIND and WashU-MDC cohorts employed a cross-sectional study design. A total of 2,628 participants of European descent were included in the main analysis, consisting of 1,625 PD cases (PPMI=534, PDBP=725, BioFIND=106, WashU-MDC=260) and 1,003 control participants (PPMI=141, PDBP=463, BioFIND=76, WashU-MDC=323; Table 1). Mean ages of the participants in PDBP (64.65±10.10) did not significantly differ from PPMI (63.85±9.83) (p=0.10). However, BioFIND participants, with the mean age of 67.21±6.86, were significantly older than both the PDBP (p=1.70×10^−5^) and PPMI (p=1.91×10^−7^) participants. Similarly, WashU-MDC participants, whose mean age was 69.21±9.26, were significantly older than participants in either of the other three cohorts, PDBP (p<2.20×10^−16^), PPMI (p<2.20×10^−16^) and BioFIND (p=1.89×10^−3^). Consistent with the well-documented higher prevalence of PD in males^22,23^, all four datasets exhibited a comparable higher male proportion (>60%) of PD participants (p=0.72). Healthy controls were balanced by sex in all datasets except PPMI, which had a significantly higher proportion of males (63.57%) than healthy control participants in PDBP (45.14%, p=2.70×10^-4^), BioFIND (47.36%, p=0.04), or WashU-MDC (43.96%, p=2.20×10^-4^). BioFIND PD participants had significantly higher UPDRS-III scores (39.07±13.23) compared to those from PPMI (24.84±13.15, p<0.001) or PDBP (25.83±13.36, p<0.001). Healthy control participants had similar mean UPDRS-III scores in PPMI (1.48±2.62) and PDBP (2.12±5.06; p=0.72). UPDRS-III scores were not available for WashU-MDC participants or BioFIND healthy control participants. Finally, PD participants in the PDBP cohort presented with significantly lower MoCA scores (26.62±2.55) compared to those in PPMI (26.36±3.08, p=8.30×10^−9^) or BioFIND (26.84±2.57, p=3.70×10^−6^). Likewise, PDBP healthy controls participants had significantly lower MoCA scores (26.62±2.55) than either PPMI (26.62±2.55, p=3.80×10^−3^) or BioFIND (27.99±1.52, p=2.40×10^−5^) healthy control participants. No comparison was performed to the WashU-MDC MoCA scores, due to absence of information.

### Data processing and quality control

We accessed raw transcriptomic data (fastq files) from 7,598 ribodepleted blood RNA samples across the PDBP, PPMI, BioFIND, and WashU-MDC studies. Detailed information on sample collection and data generation for these datasets are described elsewhere.^11,20,24,25^ The publicly available datasets, PDBP, PPMI, and BioFIND, were accessed through the Accelerating Medicines Partnership Parkinson’s Disease (AMP-PD) Knowledge Portal. While all the data processing and quality control (QC) have previously been completed for each of the datasets, we processed and QC-ed the data de novo to ensure that the same criteria were applied to all datasets. We processed the raw data using in-house pipelines to generate gene-level expression counts.^20^ Briefly, we aligned the raw reads to the human genome reference (GRCh38) and annotated transcripts with Gencode 33 annotation using STAR (v.2.7.1a).^26^ Alignment quality was assessed via Picard metrics (v.2.8.2)^27^and gene expression quantified with Salmon (v.1.2.0)^28^, collapsing transcripts to gene level. To maintain high quality standards, we removed from downstream analyses any samples with low mapping rates (<50% of reads mapped in STAR or Salmon), those that failed QC in more than four categories defined by FastQC, or that presented outliers in the Principal Component Analyses (PCA). PCA outliers were deemed those samples whose values of the first or second principal component differed more than three standard deviations from the mean values of those respective principal components. Additionally, transcripts with low expression, described as having fewer than 10 reads in >90% of individuals, were excluded from analyses. Normalization was performed using the variance stabilizing transformation (vst) function from the DESeq2^29^ package to adjust for library complexity and generate the count matrices used in PCA.

### Differential abundance analysis and pathway analyses

Differential expression (DE) analyses were performed using DESeq2.^29^ Analyses were carried out on gene-collapsed transcript counts and adjusted by participants’ age at blood collection and sex as well as relevant technical variables.^30^ To account for major confounding factors in blood transcriptomics, such as variation in cellular composition, cell proportions were obtained using deconvolute function from the immunedeconv^31^ package and subsequently included in the model. Results of DE analyses from all four cohorts were integrated via meta-analysis using the metaRNAseq^33^ package. Only genes exhibiting the same direction of effect across all four datasets were considered for the meta-analysis. Benjamini-Hochberg adjusted p-values lower than 0.05 were considered statistically significant. We performed pathway analyses on the DE genes identified in the meta-analysis using ClusterProfiler^34^ and DOSE^35^ packages.

### Sensitivity analyses

To examine if the findings were independent of ancestry, we also assessed transcriptomic patterns in 126 participants of African ancestry (PPMI=12, PDBP=15, BioFIND=9, WashU-MDC=90; Supplementary Table 1). Due to limited sample size, data from all four cohorts was combined into a single dataset for these analyses. Prior to combining, transcript counts were scaled between the four cohorts by calculating z-scores. Normalized, scaled counts were then fit to a linear regression model to compute the difference in expression levels between PD and healthy control participants for each transcript.

Next, to determine if PD associated mutations affect the whole blood transcriptomic landscape, we investigated the expression patterns in 124 LRRK2, 39 GBA1, and 16 SNCA symptomatic participants (Supplementary Table 1) from PPMI. The most common mutations in the symptomatic participants were G2019S (n=90) in LRRK2, N409S (n=31) in GBA1 and A53T (n=16) in SNCA participants. Additionally, we explored whether the changes in expression of the DE transcripts start prior to symptom onset by assessing expression in those individuals without symptoms but at risk of developing PD, such as asymptomatic participants with PD-associated mutations (160 LRRK2 and 96 GBA1), as well as participants who exhibited REM sleep behavior disorder (RBD, n=28) or hyposmia (n=18; Supplementary Table 1). Similar to symptomatic participants, the most common mutation in at-risk LRRK2 participants was G2019S (n=119), and N409S (n=66) in GBA1 participants. Detailed mutation carrier information for symptomatic participants, as well as data for at-risk participants, was only available in the PPMI cohort.

### Correlation of DE transcripts and clinically relevant parameters

We used DESeq2 to explore the effect of PD medication on transcript expression in symptomatic participants. Detailed medication data was available for the PPMI cohort only, limiting these analyses to the 486 participants with levodopa equivalent information.

To explore the clinical relevance of the identified transcripts, we assessed correlations between normalized transcript counts and UPDRS-III or MoCA scores in participants with available data (regardless of diagnosis). We performed linear regression analysis, adjusted by participants’ age and sex. Due to data availability, these analyses were done in PDBP, PPMI and BioFIND cohorts, separately. Afterward, we employed combinePvalues function from the multiGSEA (version 1.14.0)^36^ package to calculate the combined p-value via Fisher’s combined probability test. We considered correlations statistically significant at a p-value threshold of 0.05. Further, to determine if transcript levels were causally associated with medication usage and motor symptom severity, we performed mediation analyses using R package mediation.^37^

### Multiomic data integration

To further biologically frame our results, we substantiated our findings through integration with: (i) in-house brain bulk RNAseq, and (ii) publicly available plasma and (iii) CSF proteomic data.^38^ After DE of the brain RNAseq, or the differential accumulation analyses of the two proteomic datasets, we evaluated the overlap between nominally significant findings from each of the accessed datasets and the DE transcripts identified in whole blood. We used the hypergeometric test (phyper function in R) to test the significance of each overlap. P values lower than 0.05 were considered significant.

Further, we leveraged the latest PD genome wide association studies (GWAS)^39,40^ and information compiled in the PD GWAS locus browser^41^ to query whether any of the identified DE transcripts originated from known PD associated loci. To compare our results to PD GWAS, all lead SNP coordinates were converted from hg19 to hg38 version of the human genome. Next, coordinates 500kb upstream and downstream from each SNP were found, and all overlapping regions collapsed into 69 non-overlapping genomic regions. Subsequently, bedtools intersect from the BEDTools tool suite^42^ was used to check for overlap between the collapsed PD-associated genomic regions and start and end coordinates of the identified DE genes. Finally, we browsed the PD GWAS locus browser^41^ and recorded the scores, which rank the genes based on the level of support, for those genes that overlapped PD-associated loci.

### Expression Quantitative Loci (eQTL) mapping

To identify the genetic loci that were involved in the regulation of the identified DE transcripts, we conducted a GWAS specifically on the DE transcripts to identify the eQTLs. We accessed the genotyping information from the WashU-MDC participants^18–20^ and followed standard pipelines.^20^ In brief, all samples were genotyped using Illumina GSA SNP array technology and variants were called using GenomeStudio. Imputation was performed on the TOPMed Imputation Server using the GRCh38 version R2 reference panel. Stringent quality control was applied before and after imputation. In summary, individuals or SNPs with a call rate lower than 98% were removed. Autosomal SNPs that were not in Hardy-Weinberg equilibrium (P<1×10^-06^) were also removed. The SNPs from the X chromosome were used to infer sex based on heterozygosity rates. Those samples with discordances between inferred sex and reported sex were removed. Additional QC included removal of palindromic variants, pairwise genome-wide estimates of proportion identity-by-descent to identify and remove unexpected duplicates, and removal of cryptically related samples (PI-HAT>0.30). Finally, principal component analysis (PCA) was performed using 1K genomes as an anchor to identify non-Hispanic White participants to keep the population homogeneous as possible for analyses. GWAS was conducted using PLINK2 with a linear regression model.^43^ To account for potential confounding factors, all analyses were adjusted by age, sex and genetic principal components one through five. The genome-wide significance threshold was set at p<5×10^-08^. Finally, to assess if the effect size of a genetic variant known to be associated with PD is mediated by the gene expression of any of the DE transcripts identified in here, or in other words, to infer causality between PD and the DE transcripts, we used the summary-data-based MR^35^ to test for association between the expression level of a gene and PD genetic risk using the latest PD GWAS.

### Predictive model development and evaluation

We leveraged DE transcripts and a previously established pipeline^9,44^ to construct predictive models. Briefly, vst-normalized and age and sex adjusted transcript counts were scaled using z-scores to account for technical differences between the different datasets. Next, we utilized the R package entropy v1.3.1^45^ to compute Kullback-Leibler divergence (KLD) for each transcript in the training (PDBP) compared to the testing (PPMI) dataset. Subsequently, we trained 100 L2 regularization linear models with transcripts ranging in KLD value from 0.01 to 1 in increments of 0.01 and calculated the Area Under the Receiver Operating Characteristic (ROC) Curve (AUC) value for all models in the training dataset. PDBP was used to train, PPMI to test. WashU-MDC and BioFIND were used to validate the predictive models. Additionally, and only for the PPMI dataset, we evaluated the performance of the selected models in PD participants with PD-associated mutations, as well as the ability of the models to differentiate between healthy controls and at-risk participants.

## RESULTS

### Whole blood captures PD associated transcriptomic patterns

To identify transcripts that were associated with PD, we leveraged four independent whole blood transcriptomic datasets, PDBP, PPMI, BioFIND, and WashU-MDC, totaling 1,625 non-Hispanic white PD participants (N_PDBP_=725, N_PPMI_=534, N_BioFIND_=106, N_WashU-MDC_=260) and 1,003 healthy control participants (N_PDBP_=463, N_PPMI_=141, N_BioFIND_=76, N_WashU-MDC_=323; Table 1, Figure 1). Following rigorous QC, we performed DE analyses in each population and integrated the results from all four populations via meta-analysis (Figure 1). In the meta-analysis, we evaluated 2,228 transcripts that had the same direction of effect across all four populations and found 296 transcripts to be DE when comparing PD to healthy controls, 141 of those downregulated and 155 upregulated (Figure 2A, Supplementary Figure 1, Supplementary Table 2). Additionally, six of the 296 DE transcripts, referred hereon after as the six high-confidence transcripts, NCBP3 (log_2_FC=-0.02, p=2.10×10^−9^), DLD (log_2_FC=-0.02, p=1.96×10^−7^), RNF169 (log_2_FC=0.04, p<2.20×10^−16^), CCDC57 (log_2_FC=-0.04, p=8.84×10^−13^), MRTFA (log_2_FC=-2.73, p=9.45×10^−12^), and ZKSCAN8 (log_2_FC=0.02, p<2.20×10^−16^; Supplementary Figure 1, Supplementary Table 2),were nominally significant across all four cohorts and one, RNF169, was significant after multiple test correction in each of the four cohorts.

**Figure 1.**
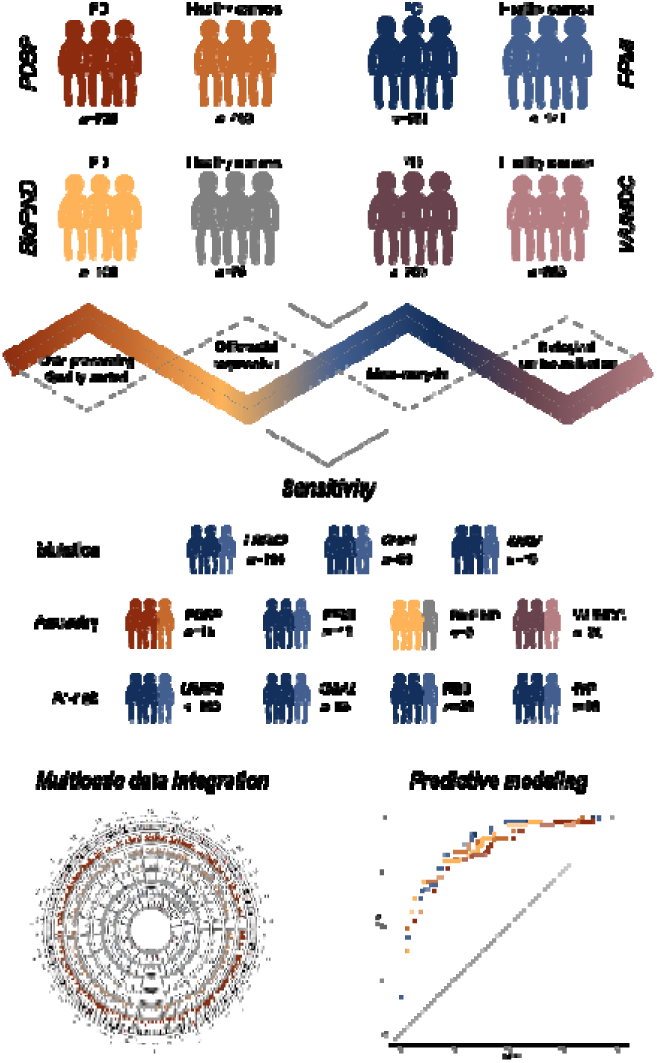
Study design.

**Figure 2.**
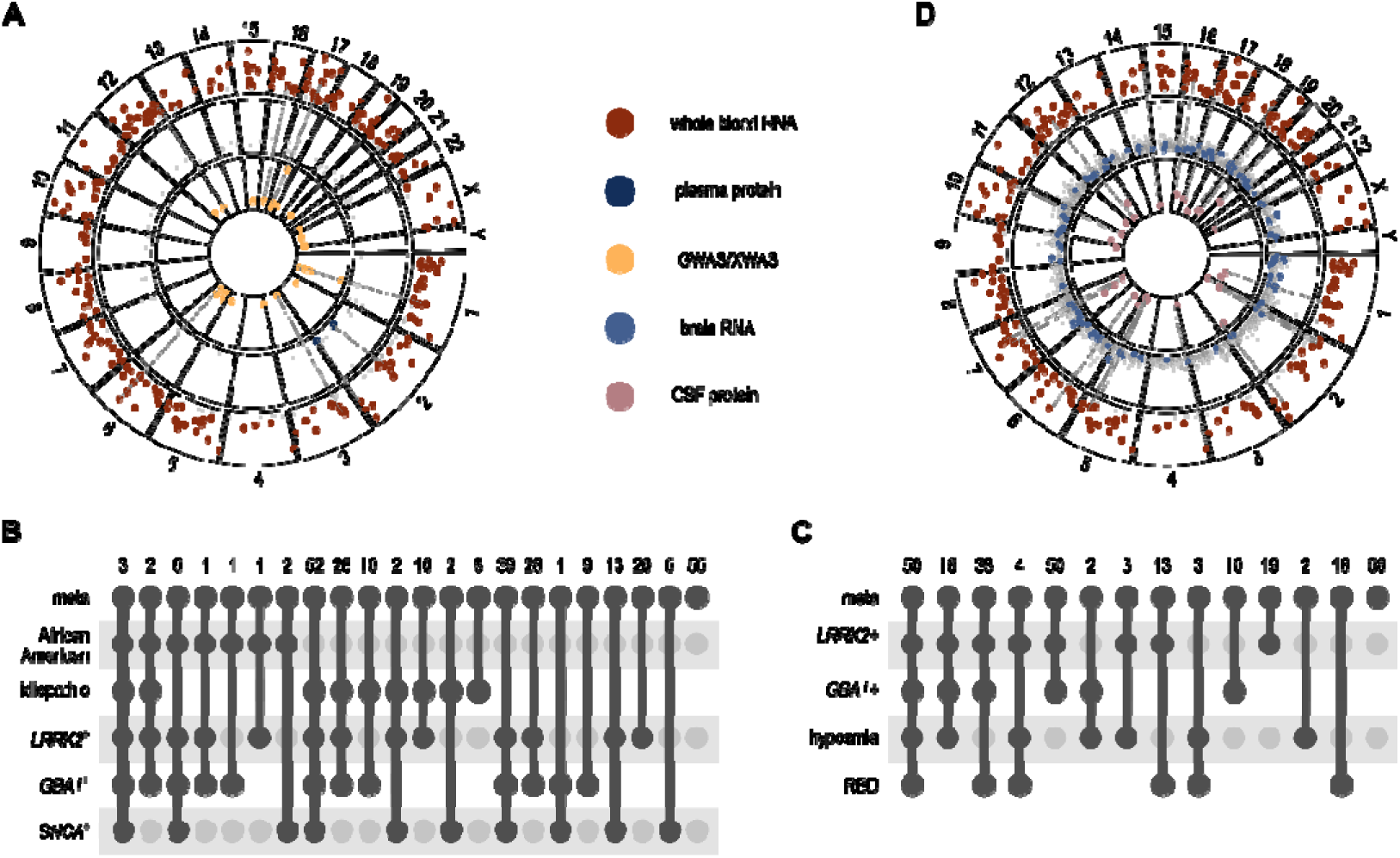
Multi-omic overlap and sensitivity analyses. A. Integration between differential expression analysis of whole blood RNAseq (maroon) with plasma proteomics (dark blue) and GWAS data (light yellow); B. intersection matrix showing results of sensitivity analyses in symptomatic participants; C. intersection matrix showing results of sensitivity analyses in at-risk participants; D. Integration between differential expression analysis of whole blood RNAseq (maroon) with brain RNAseq (light blue) and CS proteomics (rosy-brown)

To assess the potential biological relevance of the 296 DE transcripts, we explored the Kyoto Encyclopedia of Genes and Genomes (KEGG) and found that the identified transcripts were significantly enriched in the nucleocytoplasmic transport pathway (p=4.05×10⁻^7^; Supplementary Table 3), driven by NCBP2 (log_2_FC=-0.01, p<2×10⁻^16^). Consistently, Gene Ontology (GO) analysis revealed significant enrichment in nucleocytoplasmic transport (p=4.57×10⁻^7^) and RNA splicing (p=1.81×10⁻^6^; Supplementary Table 4) terms. Additionally, to test whether the change in the transcript level is also reflected in the changes in protein level, we accessed publicly available PPMI plasma proteomic data generated with Olink. We identified two proteins, PROC and SCLY, that were differentially accumulated in PD patients, and translated from two of the 296 DE transcripts (Supplementary Table 2). Although this doesn’t constitute a significant overlap (p=0.15), both proteins exhibited the same direction of effect as their corresponding transcripts in blood.

### Genetic ancestry influences the blood transcriptomic landscape

We investigated the influence of ancestry on the identified DE transcripts. We found that 166 of the 296 DE transcripts had the same direction of effect in the non-Hispanic Whites and the Black/African American participants, 16 of which were nominally significant in the Black/African American participants (Supplementary Table 5). There was no significant difference (p=0.80) in effect sizes of the 166 transcripts between the non-Hispanic Whites and the Black/African American participants. Indeed, the effect sizes show strong significant correlation (ρ=0.73, p=5.17×10^−29^, Supplementary Figure 2A). None of the six high confidence transcripts that were nominally significant across all four populations in the non-Hispanic White participants were significantly DE in the Black/African American participants, however five of them displayed the same direction of effect in the Black/African American participants, NCBP3 (effect size=-5.65×10^−3^; p=0.78), DLD (effect size=-4.05×10^−4^; p=0.98), CCDC57 (effect size=-7.45×10^−3^; p=0.71), MRTFA (effect size=-0.01, p=0.35), and ZKSCAN8 (effect size=5.10×10^−3^, p=0.79; Supplementary Table 5), as observed in the European ancestry participants.

### PD related mutations exacerbate transcriptomic differences between PD participants and healthy controls

We included all PD cases, regardless of mutation status in the previous analyses, to maximize statistical power and gain a comprehensive understanding of the transcriptomic landscape in PD. In consequence wanted to investigate the impact of mutations in LRRK2, GBA1, and SNCA on the transcriptome and discovered that mutations in each of these genes indeed contribute to already identified transcriptional changes in PD participants. We found a significant overlap between the number of transcripts that had the same direction of effect and were DE in both the meta-analysis and mutation carriers, LRRK2^+^ (203/296, p=4.47×10^−92^) and GBA1^+^ (178/296, p=4.54×10^−95^) carriers compared to SNCA^+^ carriers (125/296, p=3.21×10^−54^; Figure 2, Supplementary Table 5). Overall, we observed a strong correlation between effect sizes of DE transcripts in the meta-analysis and LRRK2 (ρ=0.86, p=3.57×10^−88^), GBA1 (ρ=0.87, p=9.60×10^−92^) and SNCA (ρ=0.80, p=7.23×10^−68^), as well as idiopathic participants (ρ=0.85, p=6.83×10^−84^, Supplementary Figure 2B-E). When we focused on the six high confidence transcripts, we observed that for all six the direction of effect in idiopathic participants, as well as mutation carriers, remained the same as in the meta-analysis. Further, two of the six, RNF169 and CCDC57, were DE in both the mutation carriers and idiopathic participants, but with a stronger association and larger effect sizes in mutation carriers (LRRK2^+^: RNF169, log_2_FC=0.12, p=1.46×10^−14^; CCDC57, log_2_FC=-0.13, p=3.49×10^-8^; GBA1^+^: RNF169, log2FC=0.14, p=3.44×10^−12^; CCDC57, log2FC=-0.22, p=3.44×10^−10^; SNCA^+^: RNF169, log_2_FC=0.19, p=4.68×10^−10^; CCDC57, log_2_FC=-0.15, p=5.73×10^−3^; Supplementary Table 5). Conversely, DLD was DE in the mutation carriers (LRRK2^+^: log_2_FC=-0.10, p=7.65×10^−7^; GBA1^+^: log_2_FC=−0.07, p=0.03; SNCA^+^: log_2_FC=−0.16, p=1.24×10^−4^), and not in the idiopathic participants (log_2_FC =-0.02, p=0.10; Supplementary Table 5). The expression patterns of the remaining three transcripts showed additional heterogeneity, with NCBP3 being DE in LRRK2^+^ (log_2_FC=-0.03, p=7.38×10^−3^) and SNCA^+^(log_2_FC=-0.07, p=0.02) carriers, MRTFA in idiopathic participants (log_2_FC=-0.03, p=0.03) and LRRK2^+^ carriers (log_2_FC=-0.06, p=1.89×10^−3^), while ZKSCAN8 was exclusively DE in SNCA carriers (log_2_FC=0.07, p=0.01; Supplementary Table 5).

### Transcriptomic perturbations can be detected prior to symptom onset

We next examined whether individuals who were at risk of developing PD displayed similar transcriptomic patterns to those observed in symptomatic participants. At risk individuals included carriers of known PD-related mutations (LRRK2, GBA1, and SNCA), as well as individuals exhibiting PD-associated syndromes like RBD or hyposmia, none of whom had symptoms of PD. This included 159 LRRK2 and 59 GBA1 mutation carriers, as well as 18 participants with RBD and 27 participants with hyposmia from the PPMI cohort. Given that only six SNCA carriers were available, analyses of this group of at-risk participants were not conducted.

We found a significant enrichment in the transcripts that were already dysregulated in LRRK2 at-risk participants (193/296, p=7.22×10^−92^). Similarly, we observed dysregulation of the DE transcripts in GBA1^+^(166/296, p=3.97×10^−72^) at-risk participants, as well as in participants with RBD (126/296, p=3.41×10^−53^) and hyposmia (80/296, p=9.57×10^−35^) (Figure 2, Supplementary Table 5). We found that the effect sizes of DE transcripts were strongly correlated between the meta-analysis and all groups of at-risk participants (LRRK2^+^: ρ=0.81, p=3.94×10^−70^; GBA1^+^: ρ=0.76, p=2.64×10^−56^; RBD: ρ=0.75, p=2.46×10^−54^; HYP: ρ=0.70, p=1.16×10^−44^, Supplementary Figure 2F-I). Consistent with our findings in symptomatic participants, we observed that DLD, RNF169, and CCDC57 differentially accumulated in LRRK2 (DLD, log_2_FC=-0.06,=4.20×10^−4^; RNF169, log_2_FC=0.09, p=6.34×10^−11^; CCDC57, log FC=-017, p=6.37×10^−14^) and GBA1^+^ (DLD, log_2_FC=-0.06, p=4.49×10^−3^; RNF169, log_2_FC=0.08, p=3.01×10^−6^; CCDC57, log_2_FC=-0.20, p=9.06×10^−14^) carriers compared to healthy controls. MRTFA, too, was DE in LRRK2^+^ (log_2_FC=-0.06, p=1.58×10^−3^) and, unlike in the symptomatic participants, in GBA1^+^(log_2_FC=-0.09, p=7.71×10^−5^; Supplementary Table 5) at-risk participants. In contrast, NCBP3 was DE in LRRK2 (log_2_FC=-0.03, p=0.04), and not in GBA1, while ZKSCAN8 was DE in GBA1 (log_2_FC=0.04, p=0.01) and not in LRRK2. Moreover, we identified three of the six high confidence transcripts, RNF169 (log_2_FC=0.11, p=2.11×10^−6^), MRTFA (log_2_FC=-0.12, p=1.37×10^−3^), and DLD (log_2_FC=-0.13, p=8.24×10^−5^), to be DE in participants with RBD, and two of the six, RNF169 (log_2_FC=0.06, p=0.03) and CCDC57 (log_2_FC=-0.17, p=6.77×10^−3^; Supplementary Table 5), to be DE in participants with hyposmia.

### Transcript expression correlates with symptom severity and prescription of PD medication

To assess the clinical significance of our findings, we explored whether the expression levels of DE transcripts related to severity of motor manifestations, captured in UPDRS-III scores, or dementia, conveyed through MoCA scores. These analyses were carried out in PDBP, PPMI and BioFIND cohorts and integrated via meta-analysis. Information on symptom severity was not available for the WashU-MDC cohort. We found that 123 of the 296 DE transcripts associated significantly with UPDRS-III (Supplementary Table 6), while 106 DE transcripts had a significant association with MoCA (Supplementary Table 6). Of the 106 DE transcripts correlated with MoCA scores, 105 also related to UPDRS-III. One of the six high confidence transcripts, RNF169, was significantly associated with both UPDRS-III (p=1.13×10^−12^; Supplementary Table 6) and MoCA scores (p=3.21×10^−11^; Supplementary Table 6). Next, we investigated if the expression of the identified transcripts was affected by medication usage. We observed 89 of the 296 identified transcripts to be significantly associated with levodopa dosage (Supplementary Table 5). Further, we found that expression of three of the six high confidence transcripts, RNF169 (log_2_FC=2.78×10^−5^, p=0.03), NCBP3 (log_2_FC=-2.31×10^−5^, p=2.86×10^−7^) and MRTFA (log_2_FC=-4.36 ×10^−5^, p=1.36×10^−11^; Supplementary Table 5), associated with medication. Moreover, 34 transcripts affiliated with medication usage also correlated with UPDRS-III scores, while 30 of the 89 medication affiliated transcripts correlated with MoCA scores. We performed mediation analyses to determine whether there was a causal relationship between transcript levels and medication efficacy. We found that two transcripts, NUP43 (estimate=-4.86×10^−4^; p=0.02) and MAML3 (estimate=-5.67×10^−4^; p=0.04), had significant average causal mediation effects.

### Whole blood transcriptomic patterns capture transcriptomic changes in brain

We integrated our findings with an in-house brain RNAseq dataset to identify whether transcriptomic changes in whole blood reflect pathologic changes in the brain. Differentially expressed brain transcripts significantly overlapped with those we identified in whole blood (80/296, p=3.60×10^−19^). Of those, 54 transcripts exhibited the same direction of effect in the two tissues, including DLD (log_2_FC=-0.29, p=0.03; Supplementary Table 2). KEGG pathway enrichment analysis revealed that the 80 transcripts were nominally enriched in PPAR signaling pathway (p=0.02, Supplementary Table 3), as well as several immunity-related pathways.

To test whether any of the DE transcripts correspond to proteins differentially accumulated in CSF we accessed publicly available PPMI CSF proteomic data generated with Somalogic. We identified nine (CD55, DLD, MGAT4B, MTPAP, NPLOC4, PDGFD, PLXNC1, POR, PSMD5) differentially accumulated proteins that were translated from nine of the 296 DE transcripts (Supplementary Table 2). While the overlap with transcriptomic changes was not significant (p=0.32), six of these proteins exhibited the same direction of effect as their corresponding transcripts in blood.

### Transcripts that are differentially expressed in blood originate from PD-associated loci

Next, we inspected if any of the 296 DE transcripts were transcribed from PD-risk loci. Our analysis revealed 28 transcripts significantly (p=1.82×10^−29^) overlapping 16 previously reported PD GWAS loci^39^ (Supplementary Table 7). Nearly all loci overlapped either one (nine loci) or two (six loci) transcripts, while one locus overlapped with five DE transcripts. Of the 26 transcripts one was concordant with the genes nominated by Nalls et al.^46^ or Kim et al.^47^, whereas 25 would nominate a different gene. Our analyses suggest that for the region chr22:41434158 the gene driving the association is MRTFA instead of either of the eight putative genes nominated (ZC3H7B, POLR3H, CSDC2, PMM1, RANGAP1, MEI1, L3MBTL2, SLC25A17). For some regions such as chr3:48711556 or chr12:132487182 several transcripts are DE, which suggests that several genes are driving the association or that the region is overall dysregulated (Supplementary Table 7). Further, we found that two DE transcripts, NAA10 and PJA1, overlapped PD-associated loci on the X chromosome^40^, which constituted a significant (p=4.89×10^−4^) overlap. Additionally, we browsed the PD GWAS Locus Browser^41^ to find if any of the DE transcripts originated from genes that had supporting evidence as PD-causal genes. We discovered that twelve DE transcripts were produced from genes with an average score of 4.75 (±1.96) (Supplementary Table 7). Moreover, of the twelve transcripts, two, BRD7 and TRAF3IP2-AS1, were also DE in brain (Supplementary Table 2).

We also performed expression quantitative trait locus (eQTL) mapping to identify genomic regions regulating the DE transcripts and to evaluate their potential causal association with PD. Out of the 296 identified transcripts, 15 were found to have genome-wide significant cis-eQTLs (p<5.00×10^−8^; Supplementary Table 8, Supplementary Figure 3). To assess their relationship with PD, we conducted two-sample MR using summary statistics from the latest PD GWAs, which revealed 13 regions nominally associated with PD. Eleven of the 13 regions were already identified by the latest PD GWAS. For example, for region chr6:32411770 our analysis suggests that BRD2 (p=0.02) is the gene driving the association instead of HLA-DRB5. Similarly, for region chr1:155070849 we found CLK2 (p=4.81×10^−5^) and SCAMP3 (p=4.81×10^−5^; Supplementary Table 8) to be the drivers of the association instead of previously nominated GBAP1. The transcripts encoded by new regions and significant in the MR analyses (MRPL42 and PIGN) represent additional genes associated with PD. None of these associations remained significant after correction for multiple testing.

### Whole blood captures PD associated transcriptomic signatures even before symptom onset

We explored if the DE transcripts can distinguish between PD and healthy control participants. The four independent RNAseq datasets were used to train (PDBP), test (PPMI) and validate (WashU-MDC and BioFIND) predictive models, following the previously developed approach.^44^ We evaluated 100 predictive models equating to 100 KLD value thresholds (between 0.01 and 1) and selected the three models that had best balanced AUC-ROC values and number of transcripts (Supplementary Figure 4). The selected models contained 228, 246 and 250 transcripts and achieved AUCs of 0.727, 0.730 and 0.733, respectively, in the testing dataset (Figure 3A, Supplementary Table 13). We found that the predictive models performed better than age and sex alone which achieved an AUC of 0.584 (0.561-0.608), in predicting PD. Further, the addition of age and sex information to the predictive models lead to a marginal improvement in their performance, increasing the AUC values to 0.736, 0.742 and 0.742, for models containing 228, 246 and 250 transcripts, respectively. The models were validated, exhibiting similar AUC values in WashU-MDC (AUC_228_=0.716, AUC_246_=0.721 and AUC_250_=0.722) and BioFIND (AUC_228_=0.680, AUC_246_=0.685 and AUC_250_=0.685; Figure 3B, Supplementary Table 13) populations. We further explored if genetic background affected predictive power of the selected models. We observed that all three models had higher predictive power in symptomatic carriers of LRRK2 mutations (AUC_228_=0.789, AUC_246_=0.793 and AUC_250_=0.794), GBA1 (AUC_228_=0.870, AUC_246_=0.878 and AUC_250_=0.885) and SNCA mutations (AUC_228_=0.856, AUC_246_=0.855 and AUC_250_=0.858), compared to idiopathic participants (AUC_228_=0.681, AUC_246_=0.684 and AUC_250_=0.687; Figure 3C, Supplementary Table 13). Finally, we inspected whether the designed models could differentiate between at-risk and healthy control participants. The model performance in at-risk LRKK2 (AUC_228_=0.698, AUC_246_=0.702 and AUC_250_=0.704) and GBA (AUC_228_=0.678, AUC_246_=0.686 and AUC_250_=0.692) mutation carriers was similar to that in idiopathic PD participants, while they achieved higher AUC values in participants with hyposmia (AUC_228_=0.820, AUC_246_=0.824 and AUC_250_=0.827) and RBD (AUC_228_=0.820, AUC_246_=0.805 and AUC_250_=0.804; Figure 3D, Supplementary Table 13).

**Figure 3.**
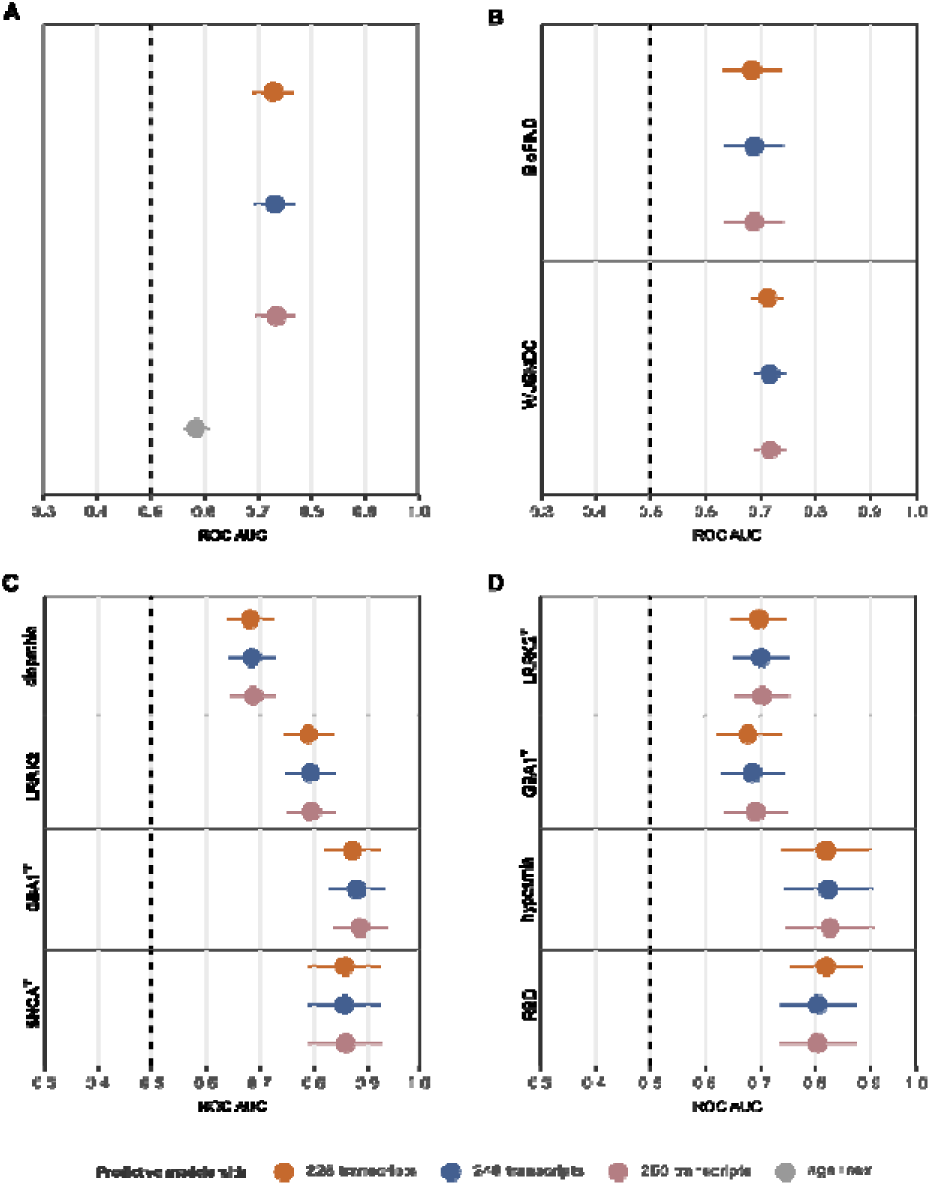
Predictive models accurately differentiate PD from healthy controls. Whisker plots showing predictive model performance in the A. testing population, B. validation populations, C. symptomatic participants stratified by mutation status, and D. at-risk participants.

## DISCUSSION

In the current study, we leveraged four large-scale whole-blood RNA-seq datasets to identify transcripts that differentially accumulated in participants with PD. To our knowledge, this represents the largest and most comprehensive PD transcriptomic analysis to date^11,13,48–51^, encompassing data from 2,628 participants across the PDBP, PPMI, BioFIND, and WashU-MDC, while also addressing distinct PD subpopulations and multi-omic data integration. We identified 296 transcripts that were DE in PD participants, many of which are implicated in pathways associated with neurodegeneration, such as nucleocytoplasmic transport.^52–54^ Furthermore, our analysis revealed that expression differences are influenced by ethnic background, highlighting the need for multi-ancestry approaches in PD research. Finally, we leveraged our findings to develop three scalable predictive models that achieved AUCs of 0.727-0.733 in the testing population. Unlike previous studies^14,51^, we used two independent cohorts to validate our models, showing that they maintain similar levels of accuracy in the validation populations. While we show that reliable, replicable predictive models can be based on transcriptomic data alone, like the previous study^14^ we suggest that the performance of the models improves by inclusion of other, non-transcriptomic features, such as genetic and clinical information, as we observed in our models with PD-associated mutations.

We found that 28 of the 296 identified transcripts originated from known PD-associated loci.^40,46,47^ Notably, two of these transcripts, STAT3 and QRICH1, scored an eight in the PD GWAS locus browser^41^, comparable to the well-established PD gene GBA1, which has a score of nine. STAT3 has been proposed as a therapeutic candidate for PD due to its neuroprotective effects. Overexpression of STAT3 has been shown to protect dopaminergic neurons from degeneration in a PD rat model.^55^ Despite not being significant in the MR analyses, we found significant correlation between STAT3 and clinical measures of motor signs (UPDRS-III) and cognitive decline (MoCA), providing further support for the involvement of STAT3. In contrast, QRICH1, while not previously related to PD, has been associated with polymicrogyria, a neurodevelopmental disorder characterized by cognitive deficits and motor abnormalities.^56^ Additionally, two of the 28 transcripts, NAA10 and PJA1, originate from loci on the X chromosome and have established roles in neurodegeneration. Given the location of these genes, further studies might investigate if they could explain, fully or partially, the higher prevalence of PD in males via dosage effect. As the catalytic subunit of the NatA complex, NAA10 is responsible for N-terminal acetylation of approximately 40-50% of all human proteins.^57–59^ This process regulates amyloid β-protein generation, modulates the stabilization of Sup35 amyloid formation, and prevents aggregation of Htt, further highlighting its importance in neurodegenerative diseases such as Parkinson’s, Alzheimer’s, and Huntington’s diseases.^60–65^ PJA1 facilitates the degradation of ataxin-3 and huntingtin polyglutamine proteins and can suppress the aggregation of FUS, SOD1, and α-synuclein in neuronal cell models.^66,67^ Together, these findings suggest that PJA1 plays a broad protective role in suppressing protein aggregation and mitigating neurodegenerative processes. Additionally, MR analyses revealed several genes nominally associated with PD and nominated novel PD-causal genes. For example, we found that instead of HLA-DRB5, BRD2 seems to be driving the association in the region chr6:32411770. Recent reports found that BRD2 expression was induced by α-synuclein, bringing upon neuroinflammation typical of PD^68^, further supporting our findings.

Transcriptional changes in blood were mirrored by altered protein accumulation in plasma. Namely, PROC and SCLY, which are translated from the identified transcripts, showed dysregulation in plasma. Elevated PROC levels have been linked to a decreased risk of dementia^69^, while SCLY mediates selenoprotein turnover, which is disrupted in PD.^70–72^ Further, we found that 80 of the 296 DE transcripts in blood were also DE in brain, supporting the notion of blood-brain barrier leakage in PD.^73^ These transcripts were also enriched in the PPAR signaling pathway, a potential therapeutic target with neuroprotective effects in PD.^74,75^ The downregulation of PSMD5, a proteolysosomal gene, which was dysregulated on both transcript level in blood and protein level in CSF, has been associated with PD pathology in myeloid cells, where it plays a role in regulating protein degradation and maintaining cellular homeostasis.^76^ In fact, protein products of nine of the 296 DE transcripts (CD55, DLD, MGAT4B, MTPAP, NPLOC4, PDGFD, PLXNC1, POR, and PSMD5) differentially accumulated in CSF, six with the same direction of effect in blood and CSF, further validating the relevance of our blood findings to PD pathobiology and nominating these proteins for further investigation as biomarkers.

In addition to being DE in blood transcriptomic and CSF proteomic data, we identified a strong cis eQTL for CD55 that was suggestive in the MR, pointing to its possible role in the pathobiology of PD. On top of that, we found that CD55 expression levels correlated significantly with UPDRS-III and MoCA scores. Finally, CD55 was included in all three predictive models. Upregulation of CD55 has been shown to inhibit complement-mediated synaptic phagocytosis in a PD mouse model.^77^ CD55, also known as decay-accelerating factor (DAF), inhibits the complement cascade, which has been implicated in the pathogenesis of PD through its association with α-synuclein and Lewy bodies. CD55 was shown to be dysregulated in PD, but not AD, further underscoring its potential role in PD-specific immune dysregulation.^78^ Furthermore, research by Wang et al. highlights a broader role for CD55 in modulating the complement system, particularly through its inhibition of the C1q-dependent complement pathway. This pathway is central to microglia-mediated synapse elimination, where CD55 disrupts complement activity and microglial phagocytosis.^79^ Altogether, this suggests that CD55 has a potential as a marker of disease progression and potential therapeutic target.

Similar to CD55, BRD7 was found to be DE in whole blood and brain tissue and mapped to a PD GWAS locus. It was further validated through eQTL mapping despite not being significant in the MR, highlighting its potentially important role. BRD7 is a bromodomain-containing protein (BCP) that has been implicated in the regulation of chromatin and gene expression. It is widely expressed in various brain regions, particularly the medial prefrontal cortex (mPFC), and localizes specifically to neurons. Inactivation of BRD7 in animal models impairs cognitive behavior and reduces synaptic plasticity, particularly in the mPFC.^80^ Additionally, BRD7 plays an anti-inflammatory role by inhibiting the NF-κB signaling pathway, a key pathway involved in neuroinflammation. BRD7 knockout mice have increased inflammation and elevated levels of inflammatory markers^81^, supporting the idea that BRD7’s normal function helps regulate neuroinflammatory processes. Moreover, recent research has shown that BRD7 complexes with Olig2, a key transcription factor, and interacts with the nucleoporin Seh1 to regulate oligodendrocyte differentiation and myelination in the central nervous system.^82^ This interaction is important for maintaining proper neural function, and myelination defects are common in neurodegenerative diseases, including PD, where the integrity of dopaminergic neurons is often compromised. These findings suggest BRD7’s central role in neuronal survival, synaptic integrity, and myelination, with potential implications for PD pathophysiology.

This study also included symptomatic participants with mutations in LRRK2, GBA1 and SNCA, as well as asymptomatic individuals at high risk for PD, such as LRRK2 and GBA1 mutation carriers or those with RBD or hyposmia. Notably, several transcripts were dysregulated before symptom onset. Among these, DLD was DE in whole blood and showed nominally significant dysregulation across all four subpopulations supporting and broadening previous reports.^83^ On top of that, analyses stratified by mutation carrier status revealed that the association between DLD expression and PD is primarily driven by PD-related mutations, with further evidence indicating that the expression of DLD in at-risk participants is already altered prior to symptom onset. Additionally, DLD was not only differentially expressed in whole blood but also exhibited differential accumulation in brain tissue and CSF and was causally linked to PD through MR analyses. Suppression of DLD has been linked to neurodegenerative disorders, including Alzheimer’s disease, where it protects against Aβ-induced pathology.^84^ Notably, genetic studies have linked the DLD locus to an increased risk of late-onset AD^85^, suggesting a potential genetic vulnerability that could be extended to other neurodegenerative conditions, including PD. DLD is a key subunit of both the alpha-ketoglutarate dehydrogenase and pyruvate dehydrogenase complexes, and reduced activity of these enzymes has been observed in post-mortem brain tissues and fibroblasts from patients with AD and PD.^86–91^ This points to a possible connection between impaired energy metabolism and neurodegeneration. Given the central role of DLD in mitochondrial energy production, its dysregulation may lead to cellular energy deficits, oxidative stress, and neuronal dysfunction—all hallmarks of PD pathology. Lastly, DLD was included in all three predictive models developed here, positioning DLD as a promising candidate for future studies as a potential early-stage biomarker or therapeutic target due to its potential causality.

The current study has several limitations. First, the sample size, despite being the largest, is still limited, especially when considering the described subgroups. In other words, despite performing sensitivity analyses to assess the impact that ancestry and genetic background have on transcriptomic landscape, the relatively small sample size of non-European ancestry populations, mutation carriers, and at-risk participants may limit the generalizability of our findings. Additionally, lack of detailed clinical and genetic information might have hampered the predictive capabilities of the predictive models. While our analyses suggest that genetic background affects the model performance, detailed mutation carrier information must be confirmed.

Limitations notwithstanding, we have robustly identified PD-associated transcripts in the largest PD transcriptomic study to date, proposed novel genes, and biomarker and therapeutic targets. We used multi-omic data integration to demonstrate that whole blood captures signatures of pathobiology. We have also developed scalable, transcript based, predictive models that can differentiate between PD and healthy controls. Moreover, by incorporating all our observations together, we identified two transcripts, CD55 and DLD, which showed great promise as highly informative early-stage biomarkers of PD. Altogether, we demonstrated that whole blood can be leveraged for development of minimally invasive biomarkers that could aid in diagnosing PD in symptomatic and asymptomatic phases.

## Supporting information

MainTable1

## Data Availability

PDBP, PPMI and BioFIND data is available from AMP-PD (https://amp-pd.org/). WashU-MDC data Summary results for all transcripts quantified across all four cohorts can be found in our transcript level browser (pdbloodtranscriptomics.WashU-MDC.edu). All original code has been deposited at GitHub (https://github.com/Ibanez-Lab/BloodLinearRNA-ParkinsonsDisease) and is publicly available as of the date of publication.

https://pdbloodtranscriptomics.WashU-MDC.edu

https://github.com/Ibanez-Lab/BloodLinearRNA-ParkinsonsDisease

## Acknowledgments

We thank all the participants and their families along with the institutions and all the staff who provided blood, without whom this study would not have been possible. This work was supported by access to equipment made possible by the Hope Center for Neurological Disorders, the Neurogenomics and Informatics Center (NGI: https://neurogenomics.WashU-MDC.edu/) and the Departments of Neurology and Psychiatry at Washington University School of Medicine.

Data used in the preparation of this article were obtained from the Accelerating Medicine Partnership® (AMP®) Parkinson’s Disease (AMP PD) Knowledge Platform. For up-to-date information on the study, visit https://www.amp-pd.org.

The AMP® PD program is a public-private partnership managed by the Foundation for the National Institutes of Health and funded by the National Institute of Neurological Disorders and Stroke (NINDS) in partnership with the Aligning Science Across Parkinson’s (ASAP) initiative; Celgene Corporation, a subsidiary of Bristol-Myers Squibb Company; GlaxoSmithKline plc (GSK); The Michael J. Fox Foundation for Parkinson’s Research; Pfizer Inc.; AbbVie Inc.; Sanofi US Services Inc.; and Verily Life Sciences.

ACCELERATING MEDICINES PARTNERSHIP and AMP are registered service marks of the U.S. Department of Health and Human Services.

Clinical data and biosamples used in preparation of this article were obtained from the (i) Michael J. Fox Foundation for Parkinson’s Research (MJFF) and National Institutes of Neurological Disorders and Stroke (NINDS) BioFIND study, (ii) NINDS Parkinson’s Disease Biomarkers Program (PDBP) and (iii) MJFF Parkinson’s Progression Markers Initiative (PPMI).

BioFIND is sponsored by The Michael J. Fox Foundation for Parkinson’s Research (MJFF) with support from the National Institute for Neurological Disorders and Stroke (NINDS). The BioFIND Investigators have not participated in reviewing the data analysis or content of the manuscript. For up-to-date information on the study, visit michaeljfox.org/news/biofind.

PPMI is sponsored by The Michael J. Fox Foundation for Parkinson’s Research and supported by a consortium of scientific partners: [list the full names of all of the PPMI funding partners found at https://www.ppmi-info.org/about-ppmi/who-we-are/study-sponsors]. The PPMI investigators have not participated in reviewing the data analysis or content of the manuscript. For up-to-date information on the study, visit www.ppmi-info.org.

The Parkinson’s Disease Biomarker Program (PDBP) consortium is supported by the National Institute of Neurological Disorders and Stroke (NINDS) at the National Institutes of Health. A full list of PDBP investigators can be found at https://pdbp.ninds.nih.gov/policy. The PDBP investigators have not participated in reviewing the data analysis or content of the manuscript.

This work was supported by grants from the Department of Defense (W81XWH2010849), Bright Focus Foundation (A2021033S), Michael J. Fox Foundation (MJFF-021599), National Institute of Health (P30AG066444, R00AG062723, U19AG03243812, R01AG053267, RF1NS075321), Alzheimer’s Association (DIAN-TU-PP-22-872356 and DIANTUOLE21725093), the American Parkinson Disease Association (APDA), the Missouri Chapter of the APDA, the Barnes-Jewish Hospital Foundation, the Paula & Rodger Riney Fund, and an NGI Pilot Grant.

