## SupplementaryFigures for "A Comprehensive Study of Circulating Blood Linear RNA nominates CD55 and DLD as novel causal genes and early-stage biomarkers for Parkinson’s Disease"

**Supplementary Figure 1.** **Differential expression analyses.** Volcano plots showing results of differential expression analyses in A. PDBP (maroon), B. PPMI (dark blue), C. BioFIND (light yellow) and 4. WashU-MDC (dark rosy-brown)

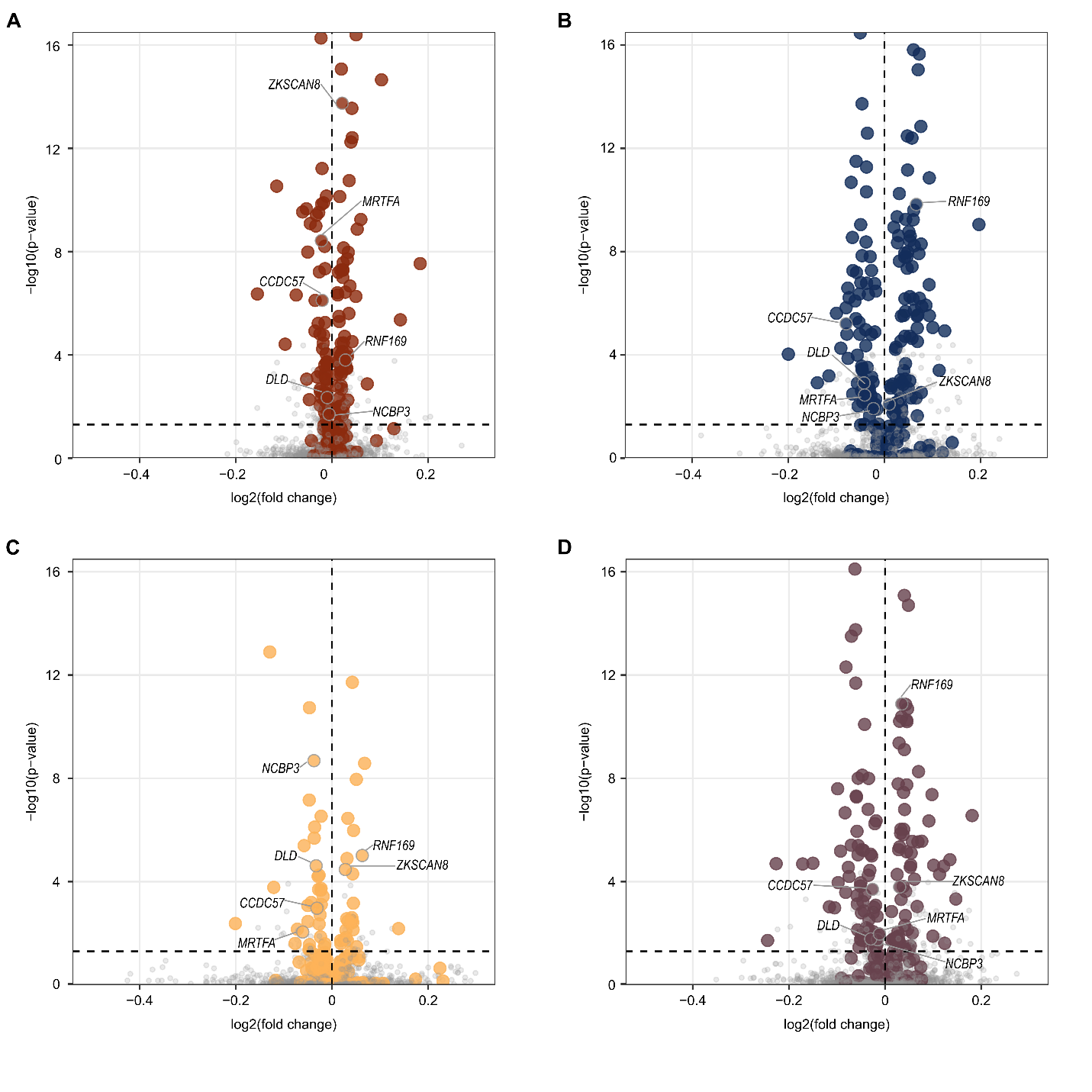

**Supplementary Figure 2. Correlation.**

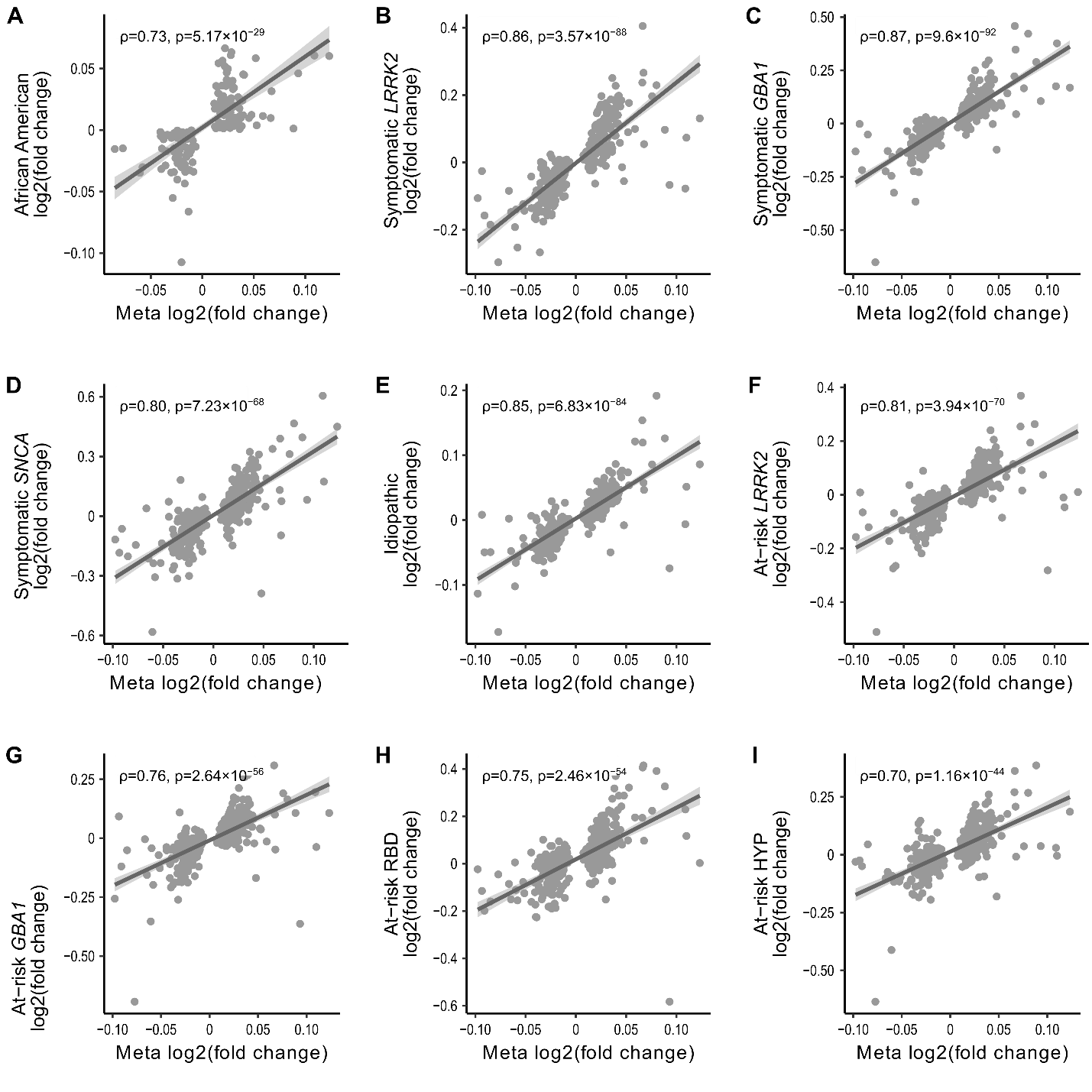

**Supplementary Figure 3. Local Manhattan plots.**

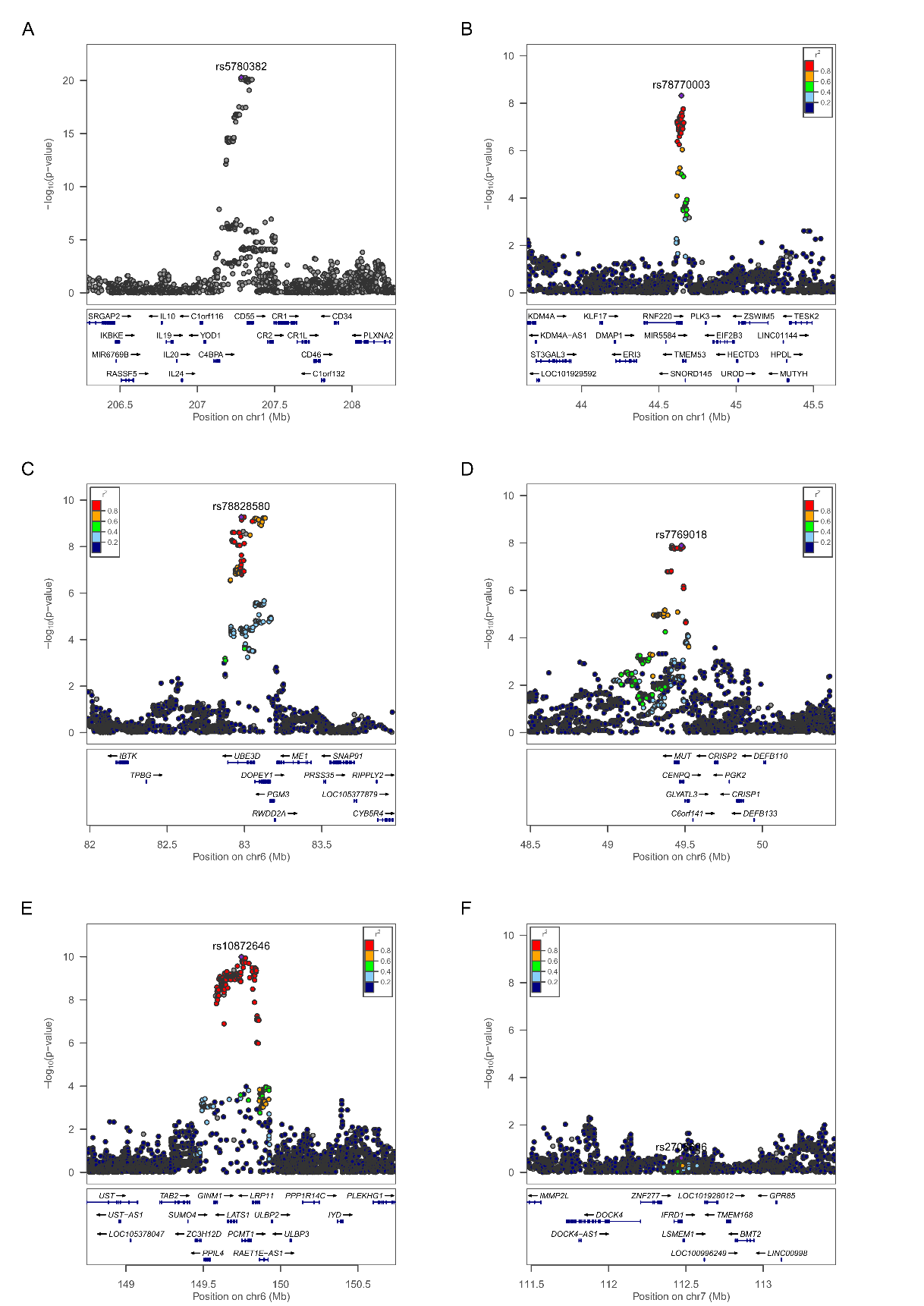

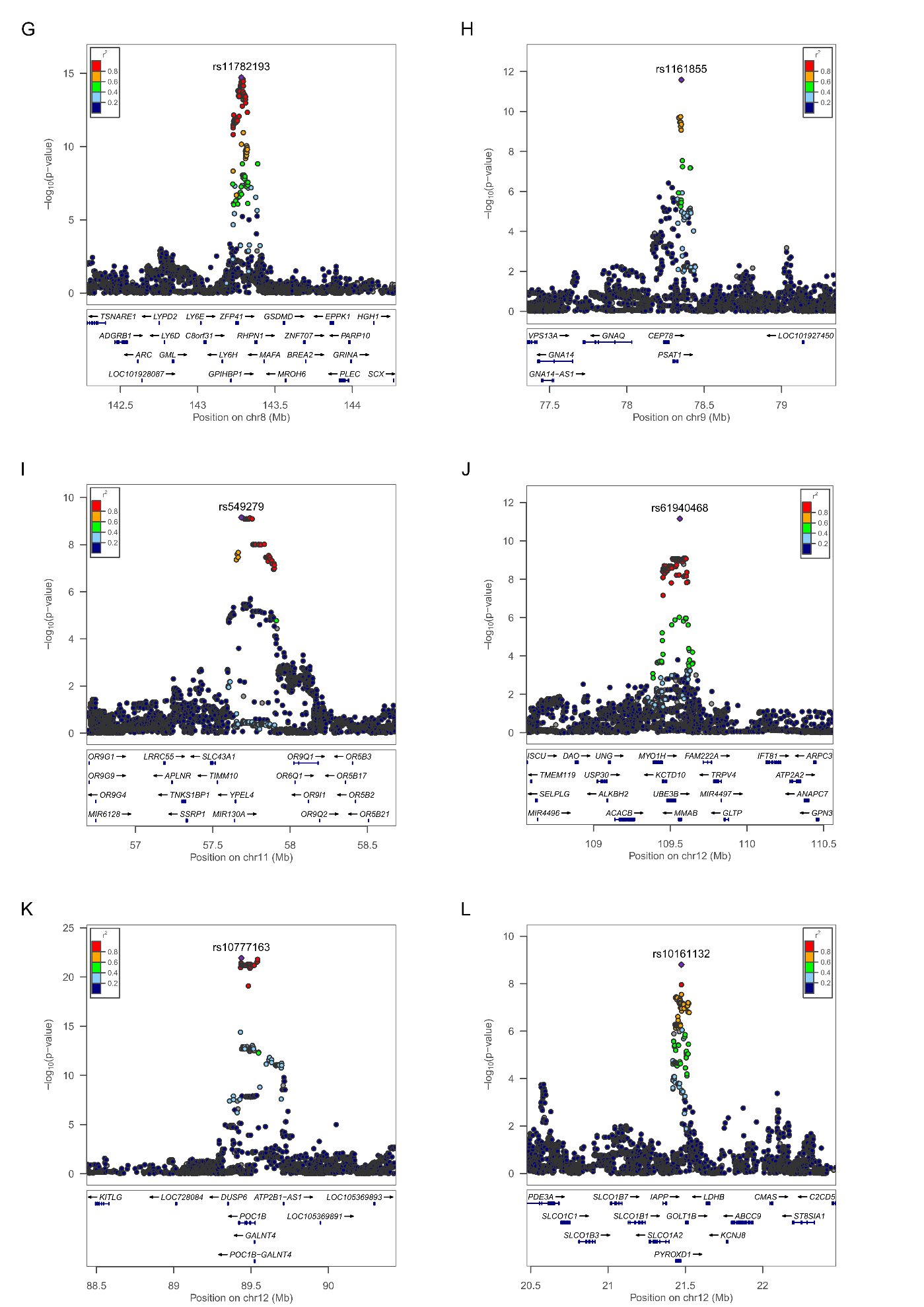

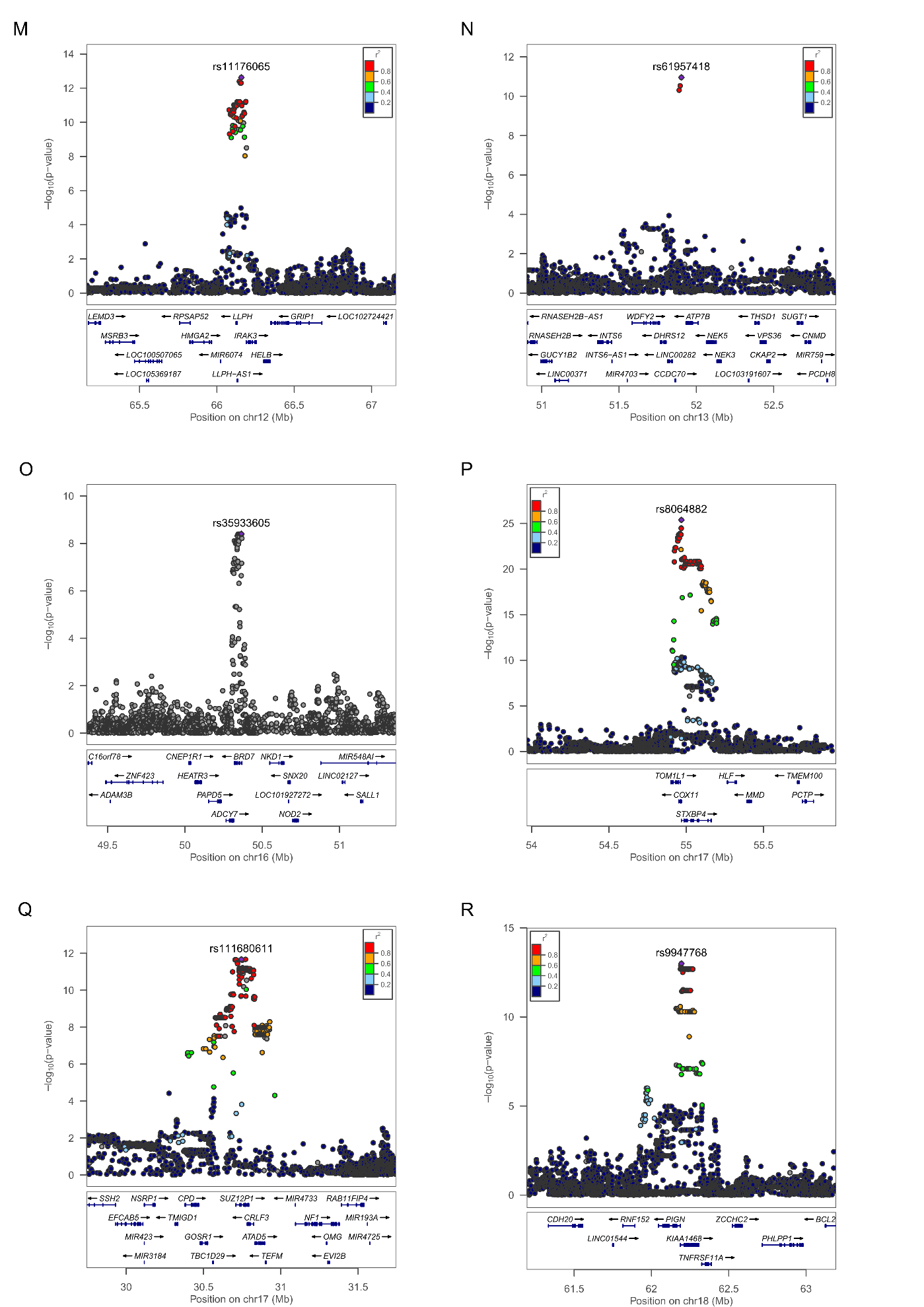

**Supplementary Figure 4.** **Feature Selection.** Model performance in the training population for increasing KLD cutoff values. The Y-axis represents the AUC of each model, and the X-axis represents the KLD value used as threshold, with models selected for further analyses highlighted in orange.

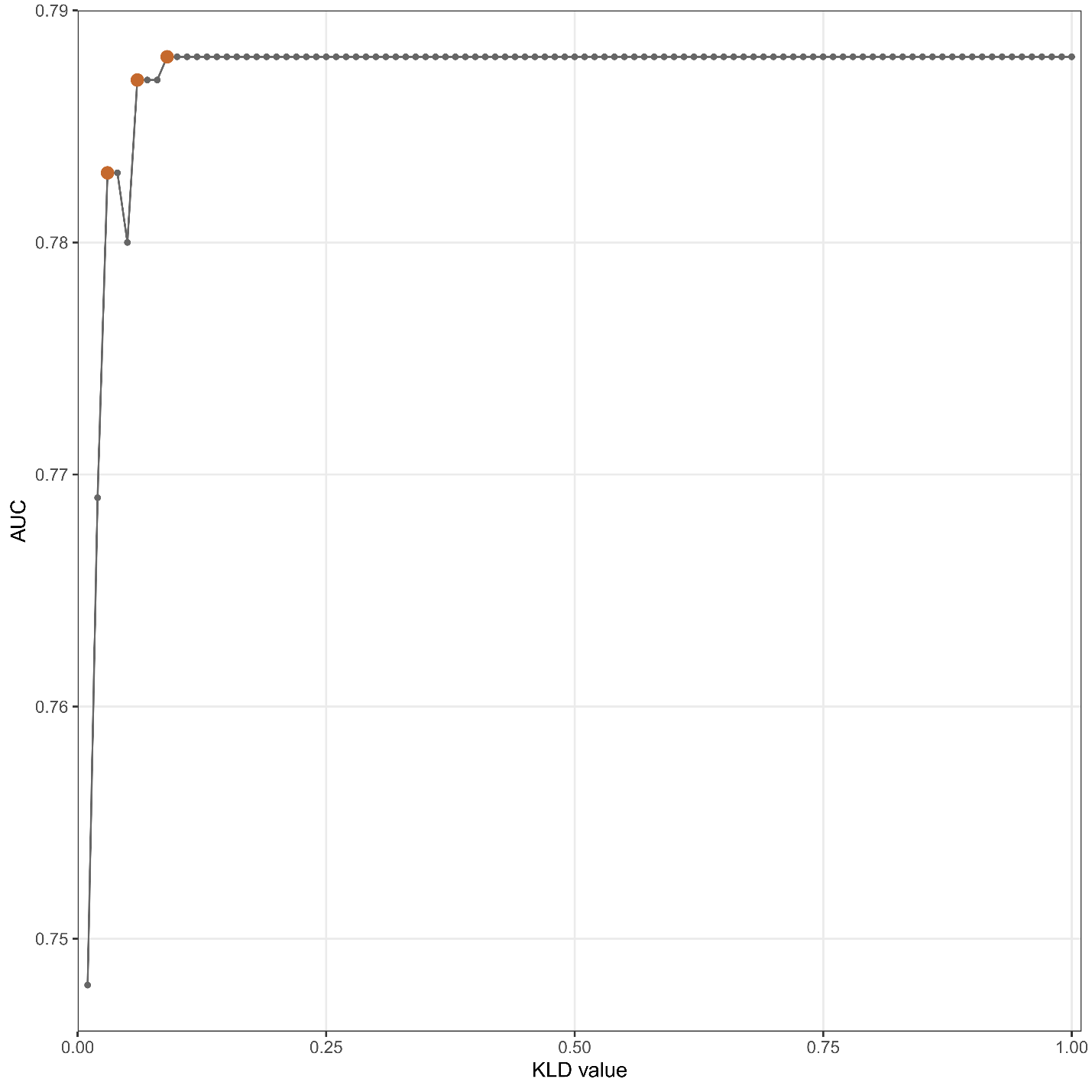
